# Developing consensus on priorities for preconception care in the general practice setting in the UK: study protocol

**DOI:** 10.1101/2024.08.26.24312573

**Authors:** Danielle Schoenaker, Elizabeth Lovegrove, Miriam Santer, Karen Matvienko-Sikar, Helen Carr, Nisreen A Alwan, Laura Kubelabo, Nathan Davies, Keith M Godfrey

## Abstract

**Background:** Preconception medical, behavioural and socioeconomic risk factors are common among people of reproductive age and can impact pregnancy and offspring outcomes. In line with clinical guidance, primary care practitioners are encouraged to support patients to manage and optimise their health prior to pregnancy. Due to barriers, including lack of time and resources, this support is not currently part of routine practice.

**Aim:** As a first step towards the co-development of practical and realistic best practice guidance, this study aims to achieve consensus on a list of priority risk factors that can be used in general practice to guide opportunistic preconception care for patients of reproductive age.

**Methods:** This study protocol was developed with a Public Advisory Group of people of reproductive age, researchers and primary care practitioners. The consensus study will consist of three steps: 1) identifying an initial long-list of candidate risk factors and defining principles for scoring the importance of each risk factor, through a literature review, workshops with people of reproductive age, and interviews with primary care practitioners; 2) stakeholder participant prioritisation of each candidate risk factor for its importance for preconception care through a three-round eDelphi survey; and 3) agreeing on the final priority list through consensus meetings with a selected group of stakeholders. Participants living in the UK will be recruited across two stakeholder groups: people of reproductive age (through the Public Advisory Group and charities) and primary care professionals (through professional organisations).

**Ethical approval:** This study has been approved by the University of Southampton Faculty of Medicine Ethics Committee (ERGO 83699 and 92950).

**Dissemination:** All study findings will be shared through stakeholder participants, peer-reviewed publication, lay summary, meetings and conference presentations, and relevant professional and community organisations. Ongoing research will inform implementation of the priority list in clinical practice.

## Introduction

Preconception care refers to a set of interventions that assess, and potentially manage and optimise, a range of medical, behavioural and social risk factors before conception in individuals of reproductive age (1). These interventions aim to improve the chance of pregnancy, pregnancy outcomes, and the health and wellbeing of parents and the next generation. Well-known preconception risk factors include, for example, lack of folic acid supplementation, smoking, obesity, domestic abuse, poorly controlled diabetes and teratogenic medication use. These factors may increase risks of infertility, miscarriage, birth defects and pre-eclampsia, as well as longer-term adverse cardiometabolic health outcomes for parents and children (2-4).

Preconception risk factors are common, with 9 in 10 women in England entering pregnancy with at least one behavioural, medical or social risk factor (5). A range of strategies – including at the population-level (e.g. mandatory flour fortification with folic acid) and the individual-level (e.g. supporting individuals planning pregnancy to optimise their health) are needed to effectively improve preconception health (6, 7). While an increasing number of population-level initiatives are underway in the UK (8-10), most individuals who conceive are not identified or supported to prepare for pregnancy and parenthood through routine universal health care.

Primary care settings, in particular general practices, are usually the first point of healthcare contact for most individuals in the UK. They are, unlike any other health service, in contact with >70% of people aged 16-49 at least once a year (11, 12) and are therefore well placed to provide preconception care. People of reproductive age and healthcare professionals agree that primary care has a key role in providing preconception care (12-15). Moreover, primary care-based interventions have been found to be effective in improving preconception risk factors, and may reduce the risk of pregnancy outcomes such as pregnancy loss and pre-eclampsia based on limited number of moderate to high quality studies to date (16).

The UK National Institute for Health and Care Excellence (NICE) Clinical Knowledge Summary on preconception advice and management (17) provides primary care practitioners with guidance on over 30 individual risk factors to assess, manage and optimise among all patients planning pregnancy, with specific additional guidance for patients with chronic physical and mental health conditions. While these assessments may take place during a dedicated preconception consultation, such consultations are not currently part of routine practice (15, 18, 19). Numerous barriers have been reported, such as limited awareness among patients and primary care practitioners, as well as lack of time, knowledge and resources (13). There is currently no guidance on best practice for the provision of preconception care in the general practice setting in the UK.

To address these barriers, primary care practitioners need practical and realistic guidance that will enable them to initiate pregnancy intention discussions with patients of reproductive age at every appropriate contact (i.e. opportunistic), screen for a limited number of key risk factors, and provide relevant advice to support optimal pregnancy planning and preparation. Such guidance will create a first step-change in the provision of opportunistic preconception care in general practice, which may be expanded as preconception care becomes more integrated and resourced, and normalised among patients and primary care practitioners.

To inform the routine provision of opportunistic preconception care, further research is needed to co-develop best practice guidance on preconception care for the general practice setting in the UK, involving relevant stakeholder groups including people of reproductive age and primary care practitioners, to ensure any barriers are identified and addressed. A first step towards the co-development of best practice guidance for preconception care is the development of an agreed list of priority risk factors that can guide primary care practitioners in providing opportunistic preconception care when time and resources are limited. This paper describes a protocol to achieve this aim.

### Study aim

The aim of this study is to reach agreement on a list of priority risk factors for adverse pregnancy and offspring health outcomes that can be used in general practice to guide opportunistic preconception care for patients of reproductive age (any gender), through a consensus process among people of reproductive age and primary care practitioners in the UK.

## Methods

We used guidance on conducting consensus studies as outlined in the Core Outcome Measures in Effectiveness Trials (COMET) handbook (20), Core Outcome Set-STAndards for Development (COS-STAD) (21) and Core Outcome Set-STAndardised Protocol (COS-STAP) (22) to design our study, where relevant.

### Scope of the preconception care priority list

The parameters of this study are informed by considerations of the target population, relevant interventions, and the context in which the priority list may be used.

#### Population

The preconception care priority list will include risk factors that are relevant to people of reproductive age (defined as 16-49 years in this study) in the UK who may conceive a child in the future, and that can be managed or optimised through routine care in the general practice setting. Informed by the NICE Clinical Knowledge Summary (17), international preconception care guidelines (23), and the latest scientific evidence (see literature search described below), risk factors may be sex-specific. For example, preconception folic acid supplementation is recommended for females trying to conceive, while interventions to address smoking and obesity may be relevant to both female and male patients (including people of all gender identities). While the aim of the consensus study is to create a simple and quick-to-implement priority list to support opportunistic preconception care, the outcome of the study may include one or two (sex-specific) priority lists. This will be informed by study findings at every step (steps described below).

#### Interventions

The preconception care priority list will inform interventions that can be provided within the general practice setting in the UK, and that may improve fertility, pregnancy outcomes, and the health and wellbeing of parents and the next generation. Preconception interventions aim to support patients to manage and/or optimise risk factors, for example through referral to stop-smoking services or social prescribing services, prescription of high-dose folic acid supplements, and assessment of cervical screening and immunisation status (17).

#### Context of use

Provision of preconception care generally starts with screening of a patient’s pregnancy intention, followed by (1) preconception intervention among patients who are considering pregnancy in the next year, (2) intervention and/or advice and education on pregnancy preparation for patients who want (more) children but not in the next year or who are unsure, or (3) advice, education and provision of contraception (if relevant) for patients who do not want (more) children (18).

We anticipate that the list of preconception risk factors agreed upon will be used as a priority list to prompt conversations with patients who are considering pregnancy in the next year, who want (more) children but not in the next year, or who are unsure. This will help guide the provision of tailored advice, education and intervention in line with the NICE Clinical Knowledge Summary (17) (or international guidelines if no NICE guidance is available for specific preconception risk factors (23)). The priority list provides an opportunity to initiate a conversation about preconception care when time is limited, while follow-up contact may be needed, especially for patients living with more complex health conditions or circumstances.

Prioritisation of risk factors that can be addressed in general practice as part of opportunistic preconception care may differ across countries, for example due to differences in national guidelines, readily available patient information, existing healthcare interventions and referral pathways, cultural differences, and population health priorities. The preconception care priority list will therefore be developed for the UK general practice context initially.

#### Study overview

This consensus study will consist of three steps (**Figure 1**):

**Figure 1.**
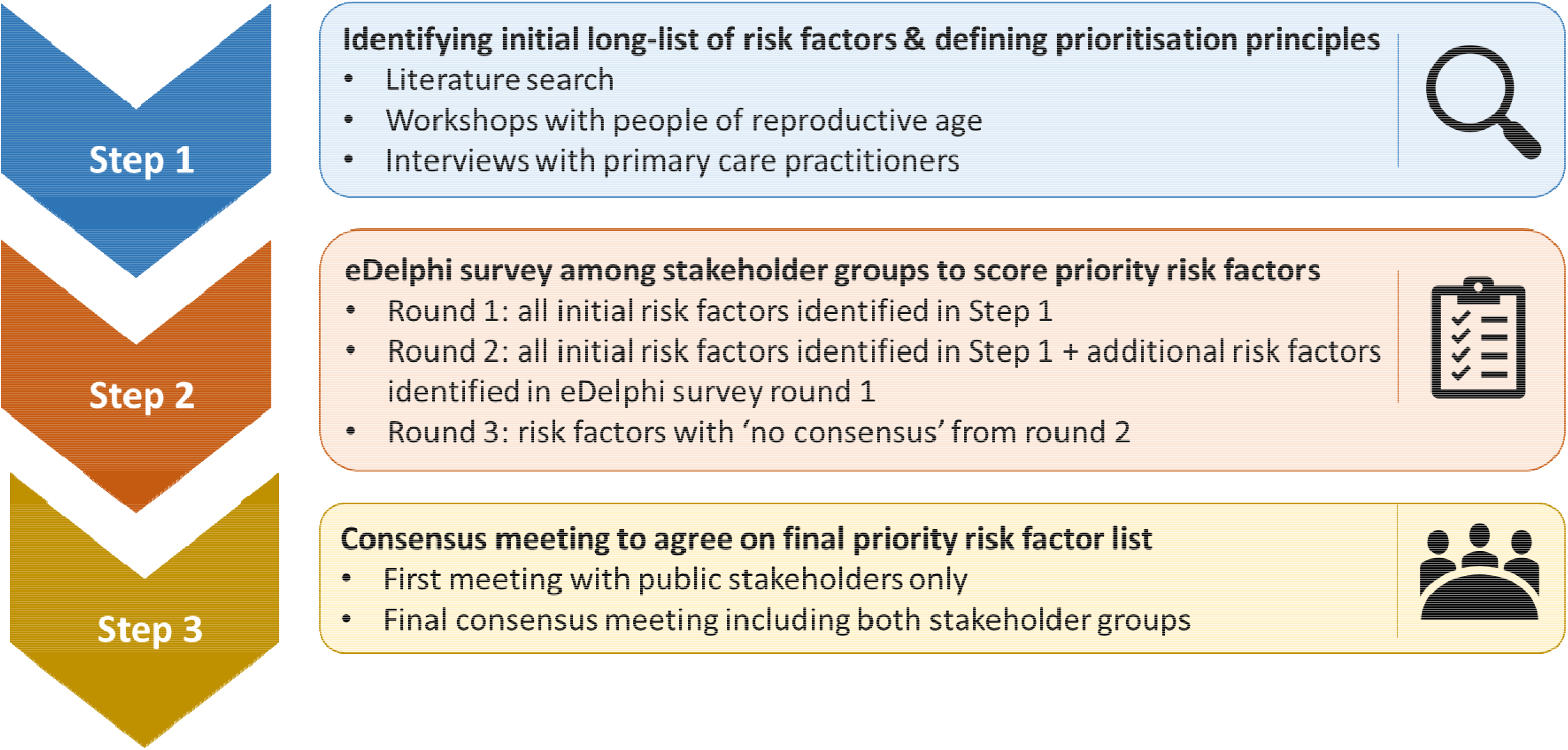
Flowchart of steps involved in reaching consensus on priority risk factors for preconception care in the general practice setting in the UK.

1. identify an initial ‘long-list’ of candidate risk factors, and define principles for scoring the importance of each risk factors, through a literature review, workshops with people of reproductive age, and interviews with healthcare professionals who care for this patient group in general practice (i.e. primary care practitioners).
2. stakeholder prioritisation of each candidate risk factor in terms of its importance for preconception care through a three-round eDelphi survey.
3. agree on the final priority list(s) through consensus meetings with a selected group of study participants.

#### Patient and public involvement (PPI)

We developed this protocol with input from a UK-wide Public Advisory Group. Through online group discussions, people of reproductive age contributed to informing the study’s PPI plan (**Figure 2**), the study aim, participant recruitment strategy and additional steps required to support people of reproductive age who take part in the study.

**Figure 2.**
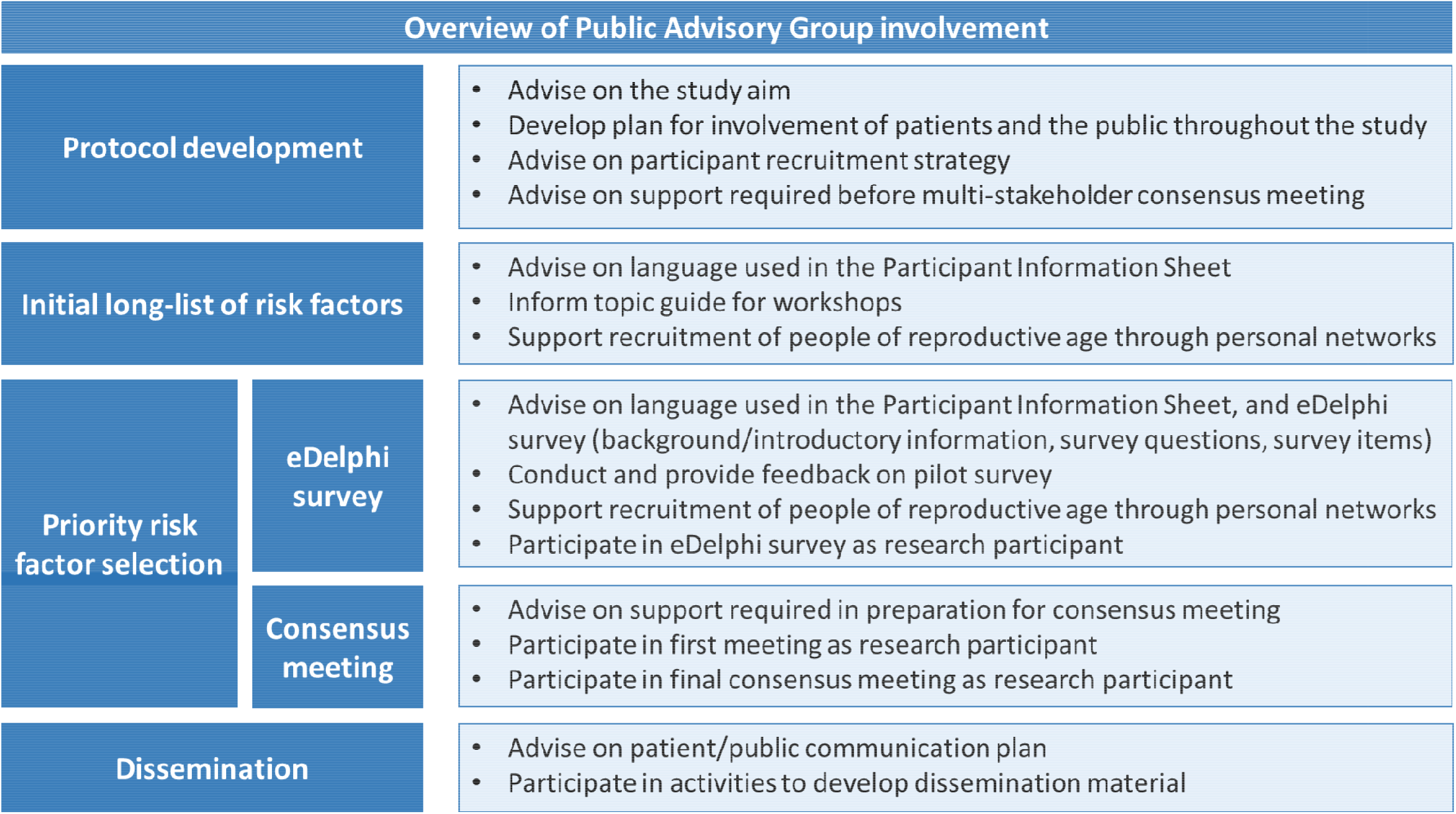
Overview of patient and public involvement in consensus study to prioritise risk factors for preconception care in the general practice setting in the UK.

Two Public Advisory Group representatives (LK and ND) have provided feedback on this protocol and the study materials. Alongside comments from the wider Public Advisory Group, they helped for example narrow down the content for the workshops with people of reproductive age (step 1), and informed the wording of the rating scale for the eDelphi survey (step 2). Throughout the study they will contribute to interpretation of results alongside the research team to inform next steps. The broader Public Advisory Group will provide feedback on study materials, pilot test the survey, support recruitment of study participants through their personal and community networks, participate in the study as research participants, and contribute to dissemination of study findings (Figure 2).

### Step 1: Identify long-list of candidate risk factors and define prioritisation principles

#### Literature search

A pragmatic approach to identifying a long-list of candidate risk factors will be used given the wide range and large number of potential preconception risk factors.

We will first identify risk factors from the NICE Clinical Knowledge Summary on preconception advice and management in line with current clinical practice recommendations in the UK (17). International preconception care guidelines may include recommendations on *additional* risk factors, which we will identify from a recent systematic review (23). Lastly, given the rapidly growing evidence in the field, we will conduct a literature search to identify umbrella reviews and systematic reviews of studies on preconception health published in English in the past 5 years to identify any *additional* preconception risk factors not currently included in national and international preconception care guidelines. We will search Medline, EMBASE and CINAHL using key search terms pre?concept* or pre?pregnan* in titles/abstracts.

Data will be extracted on: source (NICE Clinical Knowledge Summary, international guideline or umbrella/systematic review), risk factor, and population (sex/gender).

#### Workshops and interviews

We will facilitate online workshops with people of reproductive age, and online interviews with primary care practitioners working in general practice, in the UK.

The aim of these workshops and interviews is to obtain input into: (1) the list of risk factors identified from clinical guidelines and academic literature which will be circulated before the workshops/interviews (e.g. relevance, wording and grouping of risk factors); (2) additional risk factors not captured in clinical guidelines and academic literature; and (3) prioritisation principles (i.e. principles to take into account when scoring the importance of each risk factor).

Workshop information and a link to an online expression of interest form will be shared by the Public Advisory Group through social media and personal communication with their personal and community networks to recruit people of reproductive age. Primary care practitioners working with people of reproductive age in general practice will be recruited through newsletter and social media posts by the Primary Care Women’s Health Forum. We aim to obtain input from 10-20 people of reproductive age (two workshops) and 10-20 primary care practitioners (one-to-one interviews). Sampling will be purposive if a larger number of participants express interest in attending. Purposive sampling will aim for diversity in terms of age, gender and ethnicity (both groups), chronic physical and mental health conditions and frequency of contact with general practice (people of reproductive age), and type of healthcare practitioner and level of experience (primary care practitioners). Informed consent will be obtained at the time of workshop/interview invitation. Workshops will last approximately 1.5 hours and interviews approximately 30 minutes, or until no further new ideas are forthcoming. The workshops be facilitated by the lead researcher (DS) and an academic GP (EL), and interviews will be conducted by the lead researcher, based on topic guides. All sessions will be recorded via Microsoft Teams for the purpose of note-taking only (i.e. no formal qualitative analysis will be conducted). Participants will be offered a voucher in appreciation of their contribution to the study.

Collectively, the literature review, workshops and interviews will produce a long-list of candidate preconception risk factors and a list of prioritisation principles. These will be reviewed by the research team (co-authors on this paper, including GPs, researchers, and Public Advisory Group representatives), who will make final decisions on wording, grouping and inclusion of risk factors and prioritisation principles to score the importance of each risk factor for inclusion in the priority list. Members of the research team will not participate in the workshops/interviews, eDelphi survey and consensus meetings to minimise bias (20).

### Step 2: eDelphi survey

We will develop the preconception care priority list using a modified Delphi method. Delphi methodology is used to allow study participants with expert knowledge or lived experience on a particular subject to achieve convergence of opinion on the importance of different risk factors using sequential surveys (20).

#### Participant recruitment

We will invite UK-based participants from two stakeholder groups to participate in the eDelphi survey:

1. People of reproductive age will be individuals of any gender aged 16-49 years who live in the UK. They will be recruited through personal communication (including social media) by members of the project’s Public Advisory Group and their community networks, and through national charities (e.g. Sands, Beyond Reflections). Individuals who participated in the workshops and consented to being contacted about the survey will also be invited to participate.
2. Primary care practitioners may include anyone working in the general practice setting in the UK and providing care for people of reproductive age, such as GPs, practice nurses and practice-based pharmacists (24). They will be recruited through professional organisations (e.g. Primary Care Women’s Health Forum, General Practitioners Championing Perinatal Care, WiseGP, Society for Academic Primary Care). Participants will be identified through, for example, newsletters, social media or advertisement on an organisation’s webpage. Interview participants who provided consent will also be invited to complete the eDelphi survey.

Further recruitment through snowballing will be encouraged to reach a diverse range of individuals.

Participants’ stakeholder group and other characteristics (described below) will be identified through the eDelphi survey. If diversity in participant characteristics is limited, more targeted recruitment may be undertaken, however recruitment will not be based on a specific sampling frame.

#### Sample size

There is no generally accepted guidance on the optimal sample size for Delphi surveys. This is often a pragmatic choice, and not based on statistical power (20). We aim to recruit approximately 30-50 participants for each stakeholder group for the first eDelphi survey round, to account for drop-out in subsequent survey rounds. Retaining participants between survey rounds will be maximised through, for example, personalised email invitations and reminders where appropriate, including only ‘No consensus’ items based on the second round in the third survey round, and adopting a minimum time between rounds (25, 26).

#### Surveys

We will invite participants to complete three sequential rounds of the eDelphi survey. Study participants will receive the participant information sheet, have the opportunity to ask questions, and complete a consent form. They will then be able to register for the survey. Study data will be collected and managed using DelphiManager (27). DelphiManager is a secure web-based system designed specifically to facilitate the building and management of eDelphi surveys. Data will be collected on participant’s name, email address, country of residence, gender, age group and ethnicity (all participants), previous or current pregnancy, pregnancy/parenthood intentions, long-term physical and mental health conditions, frequency of contact with general practice and healthcare practitioner(s) consulted within general practice (people of reproductive age), and type of healthcare practitioner and level of experience (primary care practitioners). Participants will be given the option to enter into a prize draw to win a voucher at each survey round.

The first survey will include the long-list of all candidate risk factors identified in step 1. Participants will be asked to score the importance of each risk factor for inclusion in the priority list using a 9-point scale: ‘No or limited importance’ (scores 1-3), ‘Important but not critical’ (scores 4-6) and ‘Critically important’ (scores 7-9). An ‘Unable to score’ option will be included for each item for participants who are unclear or don’t have the knowledge to score specific risk factors. Participants will also be given the opportunity to add a maximum of five additional risk factors. A new risk factor will be incorporated into the second survey round if it is suggested by two or more participants and considered distinct from already included items based on expert opinion of the research team.

After the first survey round, data will be analysed using descriptive statistics. Responses for each risk factor will be summarised in simple bar graphs for each stakeholder group, and fed back anonymously when participants are invited to complete the second survey round. During completing of the second survey, participants will be asked to reflect on the summary scores and their own score, before deciding if they would like to keep or re-score each item. A third survey round will be conducted in a similar format as the second survey, however, only risk factors that reached no consensus in the second round based on pre-specified definitions (see below) will be included in the third survey.

If a participant does not complete the second or third survey, their scores from the most recent survey will be retained in the study. The proportion of missing responses will be reported, and the impact of attrition bias will be assessed by comparing scores from participants who completed all three rounds with those who only completed one or two rounds.

#### Consensus definition

There are numerous ways to define consensus criteria (20). For our study, it is important that the majority of participants across both stakeholder groups agree that a risk factor is of critical importance and only a small minority consider it to have no or limited importance for the risk factor to be included in the priority list. The following consensus definition will therefore be applied to identify priority risk factors:

1. ‘Consensus in’ (classified as priority risk factor): ≥70% of participants in both stakeholder groups score the risk factor ‘Critically important’ and ≤15% of participants in both stakeholder groups score the risk factor ‘No or limited importance’
2. ‘Consensus out’ (do not classify as priority risk factor): ≥70% of participants in both stakeholder groups score the risk factor ‘No or limited importance’ and ≤15% of participants in both stakeholder groups score the risk factor ‘Critically important’
3. ‘No consensus’ (for further discussion in final consensus meeting): anything else

At this stage of the study, we are not aiming for a pre-defined number of agreed risk factors, however, the research team will review the consensus definition after the first and second survey round if they consider the number of risk factors scored as ‘Consensus in’ to be too small or too large. Consensus definitions, such as the percentage cut-off of participants who need to score the risk factor ‘Critically important’, may be adjusted.

### Step 3: Consensus meetings

We will hold two consensus meetings, a first meeting with people of reproductive age only, and a final consensus meeting including both stakeholder groups to discuss the results of the eDelphi survey and reach consensus on priority risk factors for preconception care in general practice. At the end of the third eDelphi survey, all participants will be asked if they are willing to take part in the consensus meetings. Purposive sampling (described before) will be used if a larger number of participants express interest in attending. Participants will be invited to the consensus meetings after the analysis of the third survey round has been completed.

#### First meeting with people of reproductive age

A first meeting will take place only with people of reproductive age, and will aim to allow this stakeholder group to have equal opportunity to form and express their opinion (20). Based on feedback from the project’s Public Advisory Group, a separate meeting would help them to practice with Microsoft Teams functions, get to know each other, know that there will be others who will contribute to the discussion, and feel comfortable sharing opinions that will be respected. The lead researcher will facilitate one or two online meetings using a similar topic guide (5-10 participants in a meeting). Following these meetings, 5-8 people of reproductive age will be invited to participate in the final consensus meeting while considering diversity and their availability to attend the final meeting. Meeting participants will be offered a voucher in appreciation of their contribution.

#### Final consensus meeting

The final consensus meeting will bring participants from both stakeholder groups together to discuss and vote on the risk factors to be included in the final preconception priority list. This online meeting is expected to last 2-3 hours and will include 5-8 people of reproductive age and 5-8 primary care practitioners (10-16 meeting participants) to allow for meaningful engagement (20). Prior to the meeting, participants will be sent the summary scores (overall and stratified by stakeholder group) for each risk factor, and informed consent will be taken. The meeting will be chaired by a non-voting independent experienced facilitator (i.e. someone outside the research team).

First, risk factors that were scored as ‘Consensus in’ will be presented, followed by risk factors scored as ‘Consensus out’. Participants will be asked if they disagree with any of these eDelphi survey results. If there is disagreement, this will be discussed and participants will vote anonymously using a 3-point scale (‘No or limited importance’, ‘Important but not critical’, ‘Critically important’) in Mentimeter (28). If consensus is not reached following this rating, there will be further discussion and another round of rating. Second, summary scores will be presented for risk factors for which ‘No consensus’ was reached. Nominal group technique will be used to discuss these risk factors (20). Participants will be randomly allocated into small groups to discuss the ‘No consensus’ risk factors. All participants are then brought together, and each participant will be asked to express their opinion on risk factors considered for inclusion in the final preconception care priority list. This will be followed by an anonymous vote using the 3-point scale for each risk factor. Depending on the number of risk factors to be discussed, the discussions and final voting may be structured based on groups of related risk factors. These two steps will then be repeated if needed, and participants will give a final yes/no vote for each risk factor.

All risk factors that meet the ‘Consensus in’ criteria based on the eDelphi survey or receive ≥70% of votes during the meeting will be included in the priority list. A discussion will inform if risk factors can be combined or should be re-named, and if sex-specific lists are needed. If the majority of meeting participants have the opinion that the list includes too many risk factors, further ad hoc voting may take place to rank risk factors and reach agreement on the final preconception care priority list.

The meeting will be audio-recorded and transcribed using Microsoft Teams to contextualise decision making towards the final preconception priority list. Meeting participants will be offered a voucher in appreciation of their contribution.

### Ethical approval

Ethical approval for the workshops, interviews, eDelphi survey and consensus meetings has been granted from the University of Southampton Faculty of Medicine Ethics Committee and the University of Southampton Research Integrity and Governance Team (ERGO 83699 and 92950)

### Status and timeline of the study

A tentative timeline for the study is outlined in **Table 1**. At the time of manuscript submission, ethical approval for the full study was granted.

**Table 1.**
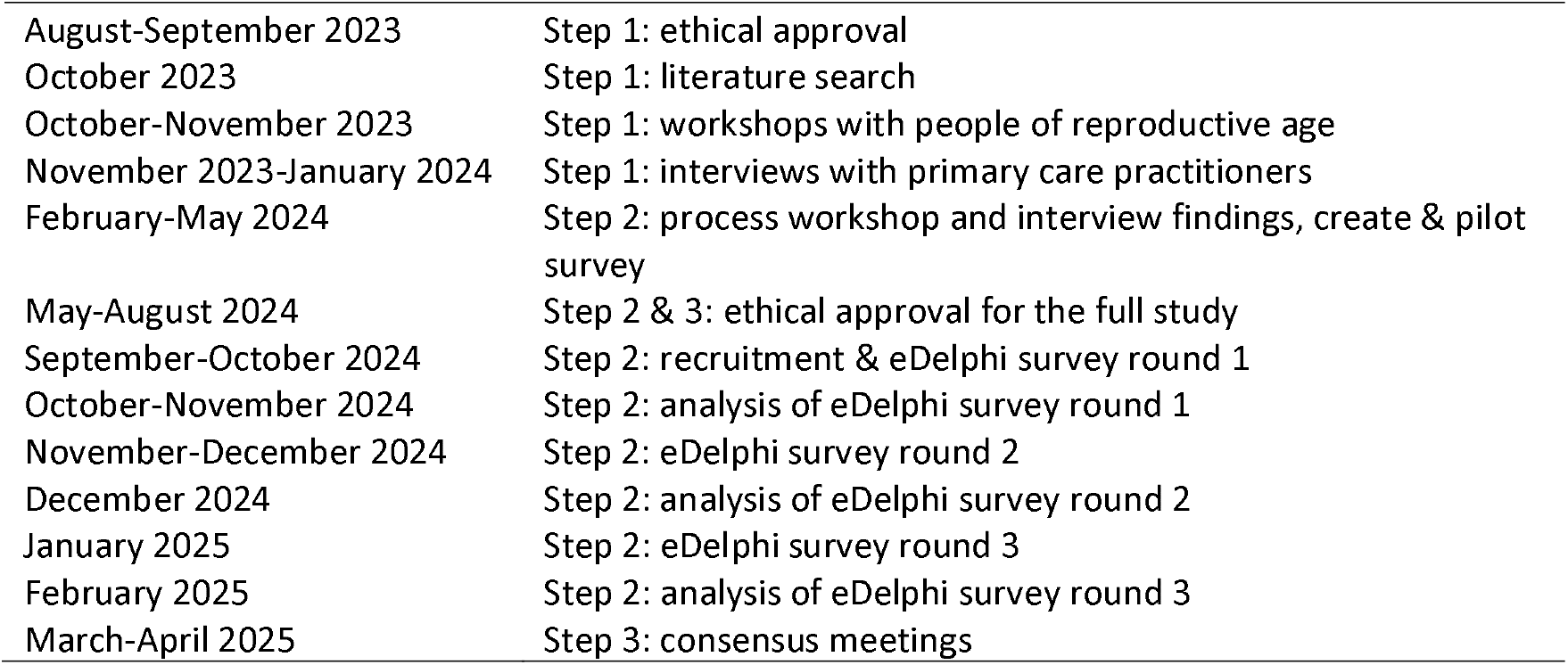
Tentative timeline of the consensus study.

|  |  |
| --- | --- |
| August-September 2023 | Step 1: ethical approval |
| October 2023 | Step 1: literature search |
| October-November 2023 | Step 1: workshops with people of reproductive age |
| November 2023-January 2024 | Step 1: interviews with primary care practitioners |
| February-May 2024 | Step 2: process workshop and interview findings, create & pilot survey |
| May-August 2024 | Step 2 & 3: ethical approval for the full study |
| September-October 2024 | Step 2: recruitment & eDelphi survey round 1 |
| October-November 2024 | Step 2: analysis of eDelphi survey round 1 |
| November-December 2024 | Step 2: eDelphi survey round 2 |
| December 2024 | Step 2: analysis of eDelphi survey round 2 |
| January 2025 | Step 2: eDelphi survey round 3 |
| February 2025 | Step 2: analysis of eDelphi survey round 3 |
| March-April 2025 | Step 3: consensus meetings |

## Discussion

This protocol describes the comprehensive methodology that will be used to conduct the first consensus study to identify the most important preconception risk factors to be assessed as part of opportunistic preconception care in primary care. Primary care has consistently been found to be among the most acceptable healthcare settings for provision of preconception care (12-15), but there is currently no co-developed practical and realistic guidance to support primary care practitioners. This study will address previously identified barriers, including lack of time, knowledge and resources (13, 19), to provide guidance for the integration of opportunistic preconception care into routine clinical practice consultations.

### Strengths

An important strength of the study is the involvement of relevant stakeholders in the design of the study, including members of the public of reproductive age and GPs. This identified, for example, a range of channels for participant recruitment including multiple relevant professional organisations, charities and community groups. We anticipate this will support recruitment of diverse members of the general practice team who may be in contact with patients of reproductive age and have a role in the provision of opportunistic preconception care, as well as diverse members of the public who contribute views based on various backgrounds and experiences.

Our study uses methodology in line with recommendations for conducting consensus studies, including three steps to reach agreement among relevant stakeholder groups. In the first step, we will review recent literature on preconception risk factors to complement risk factors that are currently included and considered in clinical guidance. Moreover, we will consult members of the public and primary care practitioners to provide their views on risk factors that may be important and relevant, but that have not been considered in the literature or clinical guidance. The comprehensive long-list of preconception risk factors will then be used in the eDelphi survey. The survey will include three rounds to give participants the opportunity to review scores from other participants on the long-list of risk factors (round 2) and on a potentially reduced list of risk factors for which study participants have not agreed yet whether they are critically important or not (round 3). Informed by input from the Public Advisory Group, a first online meeting with members of the public will be held before the final consensus meeting. Together with the use of nominal group technique, this will encourage equal participation of all stakeholders.

### Limitations

Our recruitment strategy aims to achieve representation of diverse study participants throughout the study, but it is likely we will not be able to reach all relevant groups. The workshops, interviews, surveys and consensus meetings will be conducted in English only. While people for whom English is not their first language will be able to participate, those who cannot speak or read English will not be able to take part. The use of online platforms may also exclude some participants and lead to underrepresentation of digitally disadvantaged groups. Many potential participants may also have busy work and life schedules and find it difficult to find time to participate. To address this potential barrier, the workshops and interviews will be held at varying and flexible times (including during lunch breaks and in the evening), and the survey can be completed at any time and participants can take a break and answer any remaining questions at a later stage. Moreover, a meeting poll will be completed by participants who are interested in attending the consensus meeting to find a time that suits most people.

The initial list of preconception risk factors will likely include a large number of factors, and the majority of these may be considered important for preconception care. If very few risk factors are scored ‘Consensus out’ in the first survey, then the research team will review and potentially update the consensus definition. If there is disagreement between the two stakeholder groups on the importance of many risk factors, then a large number of risk factors may need to be discussed during the final consensus meeting. Also, the importance of risk factors may largely depend on an individual patient’s health status, health behaviours and circumstances, and therefore differ for most patients. The priority list will be developed with the aim to guide an initial discussion between a primary care practitioner and a patient of reproductive age who may consider pregnancy, and further preconception care advice or consultations may be needed. This will be clarified throughout the study, and prioritisation criteria will be developed in the first step of the study to reflect this.

### Dissemination plan and next steps

All study findings will be published in peer-reviewed academic publication(s), and lay summaries, infographics, policy briefs and presentations may be developed for more targeted dissemination. These outputs will be shared with study participants (who will be encouraged to disseminate the findings to other individuals and organisations who may have an interest in the results), and relevant professional and community organisations. A detailed dissemination strategy will be developed with ongoing guidance from the Project Advisory Group and Public Advisory group.

The agreed priority list of preconception risk factors will serve to guide the provision of opportunistic preconception care for primary care practitioners to support patients of reproductive age to plan and prepare for a future pregnancy. This is the first step towards the co-development of best practice guidance that aims to meet the needs of healthcare professionals as well as patients. Ongoing research will inform how, when and by whom discussions about pregnancy and parenthood intentions and preconception advice can be best initiated, and further practical guidance and resources will be co-developed, to support the implementation of opportunistic preconception care in routine clinical practice.

## Data Availability

All data produced in the present work are contained in the manuscript.

## Acknowledgements

DS is supported by the National Institute for Health and Care Research (NIHR) through an NIHR Advanced Fellowship (NIHR302955) and the NIHR Southampton Biomedical Research Centre (NIHR203319). KMG is supported by the UK Medical Research Council (MC_UU_12011/4), the NIHR (NIHR Senior Investigator (NF-SI-0515-10042) and NIHR Southampton Biomedical Research Centre (NIHR203319)) and a Health Research Board award (HRB-EIA-2022-005). EL is supported by an NIHR In Practice Fellowship (NIHR303515). The authors would like to thank members of the Project Advisory Group and Public Advisory Group for their contributions to the development of this protocol.

## Author contributions

Conceptualisation: DS

Funding acquisition: DS

Methodology: DS, EL, MS, KM-S, HC, NAA, LK, ND, KGM

Writing – original draft: DS

Writing – review & editing: DS, EL, MS, KM-S, HC, NAA, LK, ND, KGM

## Data Availability Statement

This protocol does not report results.

